# Association of D-dimer and fibrinogen magnitude with hypercoagulability by thromboelastography in severe COVID-19

**DOI:** 10.1101/2020.07.27.20162842

**Authors:** Abhimanyu Chandel, Saloni Patolia, Mary Looby, Heidi J. Dalton, Najeebah Bade, Vikramjit Khangoora, Mehul Desai, James Lantry, Erik Osborn, Svetolik Djurkovic, Daniel Tang, Steven D. Nathan, Christopher S. King

**Affiliations:** Department of Pulmonology and Critical Care, Walter Reed National Military Medical Center, Bethesda, MD, USA; Virginia Commonwealth University School of Medicine, Richmond, VA, USA; Department of Pharmacy, Inova Fairfax Hospital, Falls Church, VA, USA; Heart and Vascular Institute, Inova Fairfax Hospital, Falls Church, VA, USA; Department of Hematology, Inova Fairfax Hospital, Falls Church, VA, USA; Department of Advanced Lung Disease and Transplant, Inova Fairfax Hospital, Falls Church, VA, USA; Department of Medical Critical Care, Inova Fairfax Hospital, Falls Church, VA, USA; Department of Cardiothoracic Surgery, Inova Fairfax Hospital, Falls Church, VA, USA

## Abstract

**Introduction:** D-dimer concentration has been used to identify candidates for intensified anticoagulant treatment for both venous thromboembolism prevention and mitigation of the microthrombotic complications associated with COVID-19. Thromboelastography (TEG) maximum amplitude (MA) has been validated as an indicator of hypercoagulability and MA ≥ 68 mm has been utilized as a marker of hypercoagulability in other conditions. We evaluated the relationship between coagulation, inflammatory, and TEG parameters in patients with COVID-19 on extracorporeal membrane oxygenation (ECMO).

**Methods:** We performed a single center retrospective analysis of consecutive patients that received ECMO for the treatment of COVID-19. TEG, inflammatory, and coagulation markers were compared in patients with and without thrombotic complications. Correlation tests were performed to identify the coagulation and inflammatory markers that best predict hypercoagulability as defined by an elevated TEG MA.

**Results:** 168 TEGs were available in 24 patients. C-reactive protein and fibrinogen were significantly higher in patients that developed a thrombotic event versus those that did not (p=0.038 and p=0.043 respectively). D-dimer was negatively correlated with TEG MA (p<0.001) while fibrinogen was positively correlated (p<0.001). A fibrinogen > 441 mg/dL had a sensitivity of 91.2% and specificity of 85.7% for the detection of MA ≥ 68 mm.

**Conclusions:** In critically ill patients with COVID-19, D-dimer concentration had an inverse relationship with hypercoagulability as measured by TEG MA. D-dimer elevation may reflect severity of COVID-19 related sepsis rather than designate patients likely to benefit from anticoagulation. Fibrinogen concentration may represent a more useful marker of hypercoagulability in this population.

## Introduction

Critically ill coronavirus disease 2019 (COVID-19) patients have been noted to display features of hypercoagulopathy involving both microthrombosis and macrothrombotic events. These patients have frequently been observed to have significant laboratory abnormalities that are compatible with a hypercoagulable state including severe derangements of D-dimer, fibrinogen, and thromboelastography (TEG) parameters.[1, 2]

Thrombosis, including venous thromboembolism (VTE), arterial thrombosis, and microthrombosis, is thought to be a sequelae of the pathogenesis of severe COVID-19 infection. Autopsies of COVID-19 infected patients have demonstrated the presence of pulmonary microthrombi and it has been postulated that microthrombus formation may contribute to end-organ dysfunction and failure.[3–5]

One approach to the management of critically ill COVID-19 patients has been the use of therapeutic anticoagulation to mitigate this hypercoagulability. Multiple local and regional organizations have advocated for the use of D-dimer testing and threshold values as a means to guide the intensity of anticoagulant therapy.[6, 7] This practice is based in part on observational evidence suggesting that treatment of patients with COVID-19 with anticoagulation is associated with improved in-hospital survival.[8] Additionally, the magnitude of D-dimer elevation independently correlates to risk of VTE in general medical patients, and furthermore, correlates to mortality in COVID-19.[9, 10]

However, D-dimer is a non-specific marker that is produced as the result of fibrin formation and degradation as a downstream product of thrombin cleavage of fibrinogen.[11] Although elevated D-dimer levels are observed with the presence of VTE, D-dimer is non-specific and often markedly elevated in other conditions including sepsis in the absence of clinically evident VTE.[12] It is not clear that the magnitude of D-dimer elevation in COVID-19 correlates with hypercoagulability or clinical conditions that are likely to benefit from anticoagulation.[2] It is also not clear that D-dimer represents a superior marker for hypercoagulability over other available tests of the coagulation system or inflammatory markers including fibrinogen, ferritin, and C-reactive protein (CRP). TEG maximum amplitude (MA), a measure of clot strength accounting for platelet and fibrin contributions, has been validated as a marker of hypercoagulability in a wide range of conditions and has been found to correlate with macrothrombotic events.[13] Previous studies have defined hypercoagulability by a TEG MA ≥ 68 mm.[14] In this study we evaluated the relationship between commonly utilized laboratory parameters and hypercoagulability as represented by TEG MA.

## Methods

We performed a retrospective medical record review of consecutive hospitalized patients admitted to Inova Fairfax Hospital, a tertiary care referral center in Falls Church, Virginia, USA between March 5, 2020 and May 29, 2020 with a diagnosis of COVID-19 confirmed by positive PCR testing and undergoing extracorporeal membrane oxygenation (ECMO). The study was approved by the Institutional Review Board (IRB# 18-3317) at Inova Fairfax Hospital.

Only patients on ECMO were evaluated as it is standard practice at our institution to obtain daily TEGs as part of the routine care of these patients. TEG was performed using the TEG 5000 platform (Haemonetics Corporation, Braintree, MA). TEGs were conducted using citrated whole blood, kaolin activator, and heparinase for patients on heparin. D-dimer values are reported by our laboratory in ug/mL of fibrinogen equivalent units (FEU).

Data was abstracted in a structured format by one of the authors (SP) and entered into a customized data form. Data was collected when available for standard TEG parameters (R-time and MA), D-dimer, fibrinogen, CRP, and ferritin. D-dimer values > 20 ug/mL FEU were recorded as 20 ug/mL FEU and R-time reported as > 60 min was denoted as 60 min. TEG and inflammatory markers or coagulation studies were compared if drawn within 24 hours. For patients on systemic heparin, TEG parameters with the use of heparin neutralization were collected. Demographic data, anticoagulant usage, and clinical outcomes including bleeding, macrothrombotic complications, and time on ECMO were recorded. Diagnosis of a macrothrombotic event (deep vein thrombosis, pulmonary embolism, or other large vessel venous or arterial blood clot) was recorded based on radiographic report finalized during hospitalization or point-of-care ultrasound findings consistent with VTE in patients with acute decompensation and suspected pulmonary embolism. Bleeding was defined as a clinically observed event, as reported by the treating clinician, that necessitated transfusion of red blood cells.

### Statistical Analysis

Distribution of all continuous data was examined for normality using visual inspection and the Wilk Shapiro test. Laboratory characteristics of the groups are presented as the median and interquartile range and compared using the Wilcoxon rank sum test. Categorical data are presented as counts with proportions and compared using Fisher’s exact test (two-tailed). A p value < 0.05 was considered statistically significant.

MA, fibrinogen, and CRP had skewed distributions. To reduce the impact of outliers and for parametric modeling, the natural log of these variables is used in graphical presentations and analyses. Associations of continuous data are presented graphically using linear regression models and plotted using natural log (ln) scales. MA values were categorized into values ≥ 68 mm and < 68 mm and the diagnostic accuracy of fibrinogen and CRP in predicting MA is presented using a receiver operator characteristic curve, summarized using the area under the curve (AUC) together with 95% CIs, and compared utilizing the empirical method. All statistical analyses were performed using STATA version 14 (StataCorp LP; College Station, TX, USA).

## Results

Twenty-four consecutive patients with COVID-19 requiring ECMO support were included in the final cohort. Patients were grouped by the presence or absence of diagnosed macrothrombosis during hospitalization. Twelve patients developed macrothrombosis either while receiving ECMO or after this therapy was discontinued (9 deep vein thromboses, 2 pulmonary emboli, and one episode of embolic mesenteric ischemia). Two patients developed bleeding complications (hemoptysis and gastrointestinal hemorrhage). Demographic data, laboratory values, and clinical outcomes between groups are displayed in Table 1. Patients with imaging confirmed macrothrombosis were significantly more likely to have higher median CRP and fibrinogen values.

**Table 1.** Characteristics of patients categorized by diagnosed macrothrombosis.

|  | **All Subjects** | **Macrothrombosis** | **No thrombosis** | **P Value** |
| --- | --- | --- | --- | --- |
|  | n=24 | n=12 | n=12 |  |
| **Age (years)** | 46 (37, 52) | 50 (43, 52) | 45 (31, 53) | 0.370 |
| **Gender, women** | 16.7 | 0 | 25 | 0.093 |
| **Race, non-white** | 83.3 | 91.7 | 75.0 | 0.590 |
| **Time prior to ECMO initiation (days)** | 5 (2, 7) | 6 (3, 8) | 5 (2, 6) | 0.400 |
| **MA (mm)** | 72.8 (71.2, 78.4) | 74.9 (72.3, 79.9) | 72.1 (70.6, 77.9) | 0.225 |
| **R time (min)** | 10.4 (8.5, 12.8) | 11.6 (8.6, 12.5) | 10.3 (8.4, 13.1) | 0.864 |
| **D-dimer (ug/mL)** | 3.5 (2.4, 7.9) | 3.1 (2.4, 4.6) | 3.9 (2.5, 9.0) | 0.312 |
| **Fibrinogen (mg/dL)** | 543.5 (476.5, 674.6) | 582.5 (538.4, 758.4) | 501.5 (447.5, 575.9) | 0.043 |
| **CRP (mg/dL)** | 11.1 (4.8, 17.8) | 15.3 (8.4, 26.9) | 7.55 (3.2, 12.3) | 0.038 |
| **Ferritin (ng/mL)** | 985.2 (631.0, 2047.9) | 1004.6 (577.8, 2258.3) | 965.8 (730.7, 1785.0) | 0.922 |
| **Bleeding complication** | 8.3 | 16.7 | 0 | 0.478 |
| **Time on ECMO (days)** | 13 (9, 20) | 16 (12, 20) | 10 (8, 16) | 0.111 |
| **Death during hospitalization** | 20.8 | 33.3 | 8.3 | 0.317 |

Data presented as median (25th percentile, 75th percentile) or n (%) unless otherwise indicated ECMO, extracorporeal membrane oxygenation; MA, maximum amplitude; CRP, C-reactive protein

All patients studied received therapeutic anticoagulation per institutional ECMO protocol with occasional cross-over between specific anticoagulant. Forty-six TEGs were obtained on heparin while 122 TEGs were obtained on bivalirudin. Matching D-dimer values were available for all TEGs and there were 7 instances where matching fibrinogen values were not obtained.

D-dimer was examined as a possible predictor of MA in patients on heparin, on bivalirudin, and in aggregate. D-dimer was found to be significantly negatively correlated with MA in all three analyses (p=0.001, <0.001, and <0.001 respectively) (Fig 1A). D-dimer explained little of the variance in MA values (adjusted R^2^=0.221, 0.112, and 0.162 respectively). Fibrinogen was found to be positively correlated with MA. Ln(fibrinogen) explained over 50% of the variance in ln(MA) (adjusted R^2^ = 0.567) and the association was significant for patients on heparin, bivalirudin, and in aggregate (p < 0.001) (Fig 1B). With ln(fibrinogen) in the model, CRP and ferritin were examined as additional predictors and were not significant.

**Fig 1.**
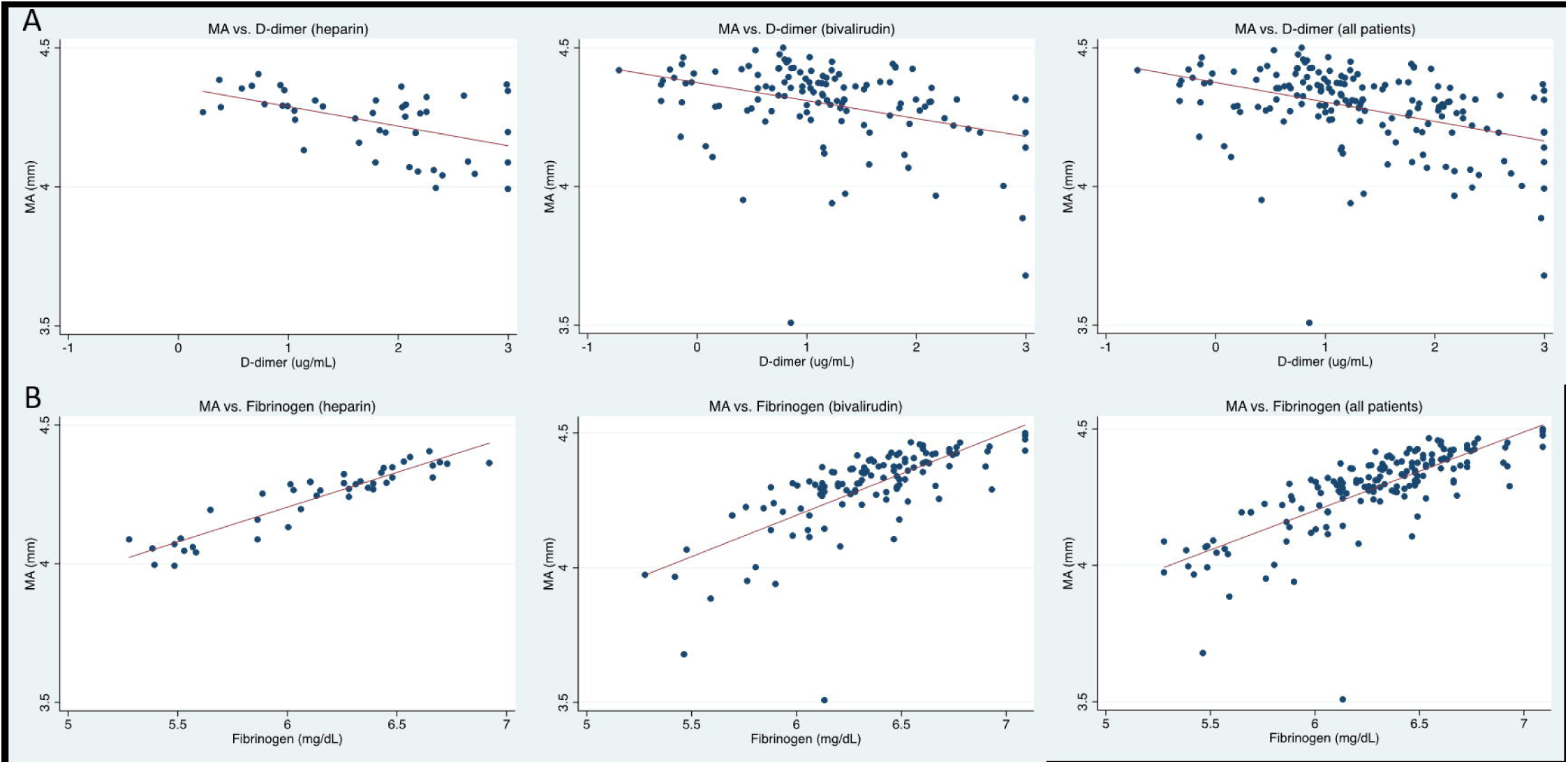
Linear regression model. (A) Ln(MA) vs. ln(D-dimer) with linear regression for patients on heparin, bivalirudin, and aggregated patients. (B) Ln(MA) vs. ln(fibrinogen) with linear regression for patients on heparin, bivalirudin, and aggregated patients.

All TEG MA values were then categorized as ≥ 68 mm and < 68 mm. MA values ≥ 68 mm were associated with lower absolute D-dimer values and higher absolute fibrinogen values (p<0.001). Receiver operator curves (ROC) based on CRP (Fig 2) and fibrinogen levels (Fig 3) were estimated. CRP accuracy in classifying MA ≥ 68 mm was poor (AUC 0.707 CI: 0.606-0.807) while classification accuracy of fibrinogen for a MA ≥ 68 mm was excellent (AUC 0.934 CI: 0.883-0.984). AUC was greater for the ROC curve utilizing fibrinogen as a predictor (p<0.001). A fibrinogen cutoff of 441 mg/dL to predict MA ≥ 68 mm (sensitivity of 91.2%, specificity 85.7%) satisfies the closest-to-(0, 1) criterion for threshold selection.

**Fig 2.**
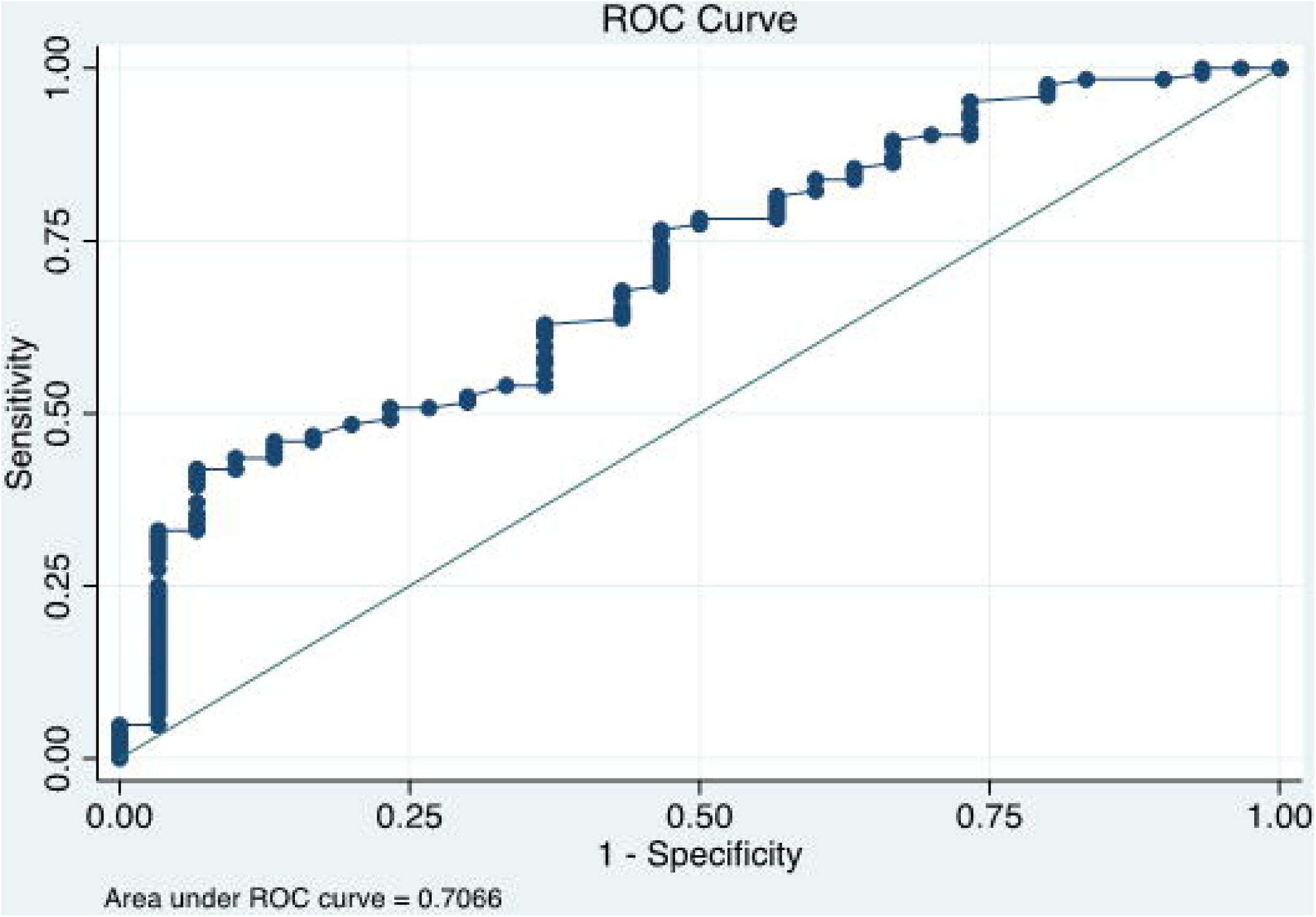
ROC curve for C-reactive protein as a predictor of MA ≥ 68 mm.

**Fig 3.**
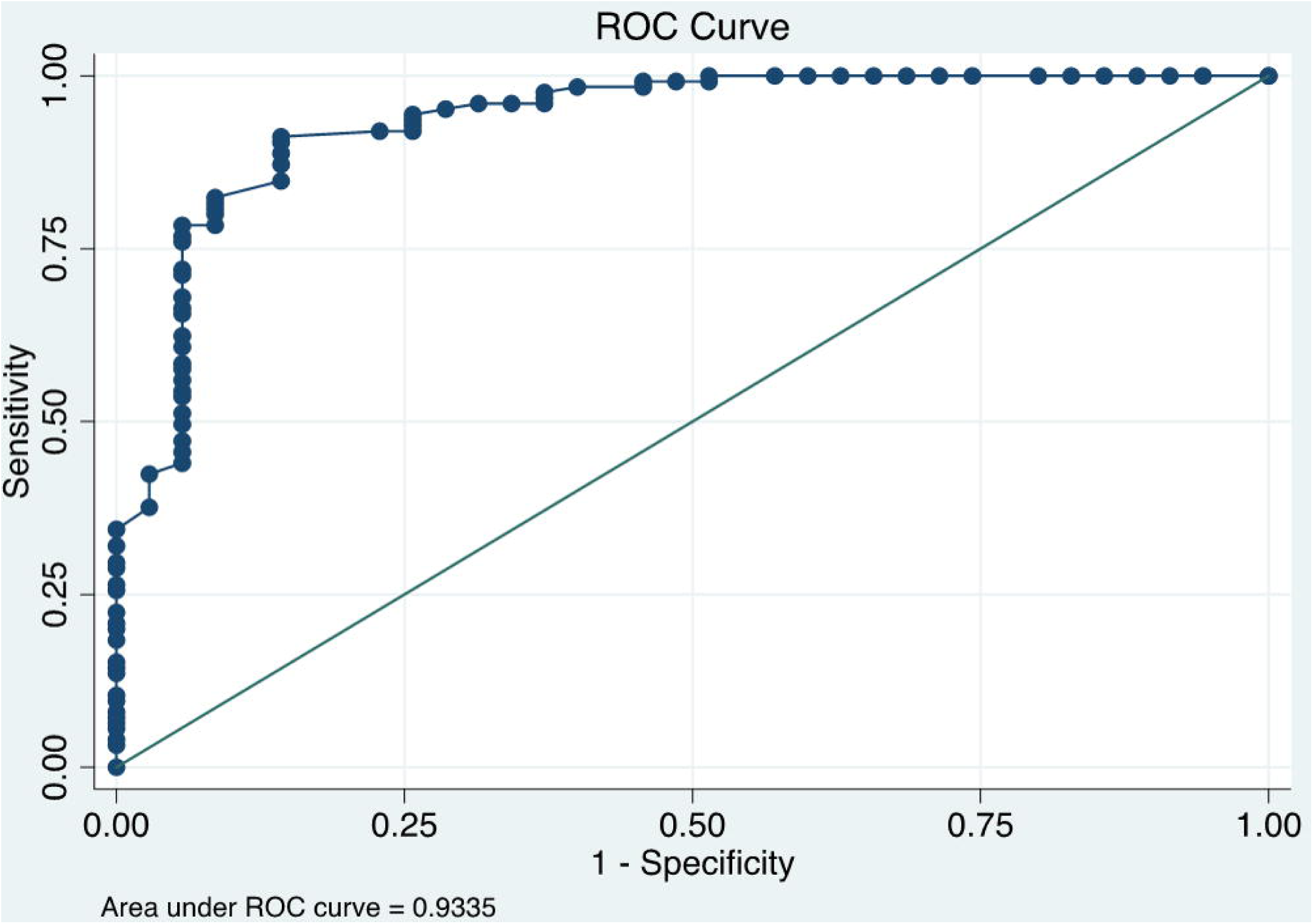
ROC curve for fibrinogen as a predictor of MA ≥ 68 mm.

## Discussion

The emergence of COVID-19 as a global pandemic has necessitated the implementation of treatment protocols based on expert opinion given limited available clinical data. D-dimer has been cited as correlating with worse clinical outcomes in COVID-19 and coagulopathy has been noted to be a common feature of severe infection.[3-5, 10] However, absolute magnitude of D-dimer has not been convincingly demonstrated to be linked to an increased risk of small or large vessel thrombosis in this disease.[2]

In this present study we aimed to evaluate the correlation of commonly available measures of coagulopathy (D-dimer and fibrinogen) with a validated reference standard (TEG). TEG MA values have previously been demonstrated to correlate with degree of hypercoagulability in a wide number of clinical conditions.[13]

Our results demonstrate that in this population of critically ill COVID-19 patients receiving ECMO therapy, absolute D-dimer magnitude did not correlate positively with degree of hypercoagulability as measured by TEG MA. On the contrary; D-dimer concentration was actually inversely related to MA in our study population. This finding was unexpected and the physiologic basis is uncertain.

A number of national guidelines have been published regarding the management of COVID-19 coagulopathy. These guidelines, in our view, are appropriately conservative with respect to recommendations related to the use of therapeutic dosing of anticoagulants for the management of critically ill patients with COVID-19.[2, 3, 15] Despite these guidelines, local and regional hospital practices vary widely, with some recommending D-dimer thresholds to aid the decision regarding initiation and intensity of anticoagulation. Based on our data, we speculate that commonly noted severe D-dimer aberrations are likely markers of COVID-19 related sepsis rather than degree of hypercoagulability. In the absence of additional data, it seems prudent to abandon the practice of D-dimer based thresholds to aid in the decision regarding the initiation of anticoagulation in COVID-19.

Studies have noted a clear association between fibrinogen and MA values in other clinical conditions.[14] Marked derangements in fibrinogen has been noted to be common in COVID-19 and an analysis of patients with COVID-19 not receiving ECMO and studied at entry into the ICU also demonstrated an association between MA and fibrinogen.[10, 16] In this study, fibrinogen levels were independent predictors of clot strength among patient with other signs of hypercoagulable status.[16] We found fibrinogen to be a much better predictor of magnitude of MA than D-dimer. In our population a fibrinogen of greater than 441 mg/dL had sensitivity of 91.2% and specificity 85.7% for detection of MA ≥ 68 mm. This finding is of interest given that TEG is not routinely available at many centers and takes specialized training to perform to achieve accurate results. Fibrinogen is a commonly available laboratory tests in most hospital environments and results may be easier to obtain. Thus, it may represent a more practical method for the identification of patients at risk for COVID-19 related hypercoagulability. A new viscoelastic device, the VCM (not yet available in the US but certified in Europe), may offer point-of-care bedside thromboelastograph results which may be useful, especially in austere environments, where fibrinogen testing is not easily available.[17]

Other investigators have compared hemostatic changes in COVID-19 to those noted in disseminated intravascular coagulation (DIC) related to sepsis. Unlike DIC; fibrinogen clotting activity in COVID-19 is frequently elevated. Further, coagulation factors such as prothrombin time and activated partial thromboplastin time do not follow the typical pattern usually seen with DIC related consumptive coagulopathy.[18] The physiologic basic of the hypercoagulable state noted in patients with COVID 19 is not well proven. We postulate that an imbalance between procoagulant factors and anticoagulant factors is logical. How elements such as factor VIII, antithrombin, von Willebrand factor, and others explain the observed hypercoagulability has yet to be fully elucidated.

Our findings should be interpreted in the context of the study limitations. First, our data is retrospectively derived from a single tertiary care center. The study was limited to severe COVID-19 infection requiring ECMO support to facilitate data collection due to the institutional use of TEG as part of the standard care of these patients. Alterations in platelet function is a known phenomenon in patients on ECMO and the implications of this alteration may affect TEG, inflammatory, and coagulation parameters.[19] Microvesicles are particles derived from platelets or monocytes and carry procoagulant activity. Microvesicles have been associated with VTE and have also been noted in patients on ECMO.[20] The interplay between COVID-19 disease and the effects of ECMO may also increase the risk of thromboembolism in these patients. Given these considerations, our selected population may limit generalizability and our findings should be validated in a non-ECMO cohort. Second, all patients in our study were on therapeutic anticoagulation with either heparin or bivalirudin per standard institutional ECMO protocol. Deviation from protocolized dosing of therapeutic anticoagulation was occasionally noted. Several of the patients in this report experienced clotting at cannulation sites despite boluses of heparin and one patient received tissue plasminogen activator in addition to anticoagulation for repetitive pulmonary emboli. There is limited data regarding the use of bivalirudin in patients with COVID-19 on ECMO and further investigation of direct thrombin inhibitors versus traditional heparin anticoagulation is needed. The role of antiplatelet agents should also be explored further in this population. As a result, the effects of anticoagulation and dosing adjustments of these medications on the study outcomes are unknown and may introduce a source of bias. Of note, all findings were consistent when examined for patients on heparin compared to patients on bivalirudin. Finally, although TEG has been validated as a useful marker of hypercoagulability; MA is a surrogate marker for more important clinical outcomes of hypercoagulability such as macro and microthrombosis. Our study lacks suitable sample size to detect the ability of MA, CRP, D-dimer, or fibrinogen to discriminate these clinical outcomes related to hypercoagulability in COVID-19. While fibrinogen and CRP demonstrated a significant increase in patients with macrothrombosis, further prospective study is necessary to better define this relationship.

## Conclusion

In critically ill patients with COVID-19, D-dimer appears to have an inverse relationship with degree of hypercoagulability as measured by TEG MA. D-dimer elevation may reflect severity of COVID-19 related sepsis rather than designate patients likely to benefit from anticoagulation. Fibrinogen concentration may represent a more useful marker of hypercoagulability in this population and further study is warranted.

## Data Availability

All supporting data will be made available after acceptance of the manuscript for
publication.

## Acknowledgments

We would like to thank all physicians, nurses, respiratory therapists, perfusionists, pharmacists, and ancillary care services who have tirelessly provided care for patients with COVID-19 within the Inova health system.

## References

1. Iba, T., et al., Coagulopathy of Coronavirus Disease 2019. Crit Care Med, 2020.

2. Moores, L.K., et al., Prevention, diagnosis and treatment of venous thromboembolism in patients with COVID-19: CHEST Guideline and Expert Panel Report. Chest, 2020.

3. Connors, J.M. and J.H. Levy, COVID-19 and its implications for thrombosis and anticoagulation. Blood, 2020. 135(23): p. 2033–2040.

4. Ackermann, M., et al., Pulmonary Vascular Endothelialitis, Thrombosis, and Angiogenesis in Covid-19. N Engl J Med, 2020.

5. Magro, C., et al., Complement associated microvascular injury and thrombosis in the pathogenesis of severe COVID-19 infection: A report of five cases. Transl Res, 2020. 220: p. 1–13.

6. Oudkerk, M., et al., Diagnosis, Prevention, and Treatment of Thromboembolic Complications in COVID-19: Report of the National Institute for Public Health of the Netherlands. Radiology, 2020: p. 201629.

7. Casini, A., et al., Thromboprophylaxis and laboratory monitoring for in-hospital patients with COVID-19 - a Swiss consensus statement by the Working Party Hemostasis. Swiss Med Wkly, 2020. 150: p. w20247.

8. Paranjpe, I., et al., Association of Treatment Dose Anticoagulation with In-Hospital Survival Among Hospitalized Patients with COVID-19. J Am Coll Cardiol, 2020.

9. Shah, K., et al., Magnitude of D-dimer matters for diagnosing pulmonary embolus. Am J Emerg Med, 2013. 31(6): p. 942–5.

10. Tang, N., et al., Abnormal coagulation parameters are associated with poor prognosis in patients with novel coronavirus pneumonia. J Thromb Haemost, 2020. 18(4): p. 844–847.

11. Adam, S.S., N.S. Key, and C.S. Greenberg, D-dimer antigen: current concepts and future prospects. Blood, 2009. 113(13): p. 2878–87.

12. Schutte, T., A. Thijs, and Y.M. Smulders, Never ignore extremely elevated D-dimer levels: they are specific for serious illness. Neth J Med, 2016. 74(10): p. 443–448.

13. Brown, W., et al., Ability of Thromboelastography to Detect Hypercoagulability: A Systematic Review and Meta-Analysis. J Orthop Trauma, 2020. 34(6): p. 278–286.

14. Kupcinskiene, K., et al., Monitoring of Hypercoagulability by Thromboelastography in Bariatric Surgery. Med Sci Monit, 2017. 23: p. 1819–1826.

15. Barnes, G., A. Cuker, and T. Gluckman, Thrombosis and COVID-19: FAQs For Current Practice. 2020.

16. Yuriditsky, E., et al., Thromboelastography Profiles of Critically Ill Patients With Coronavirus Disease 2019. Crit Care Med, 2020.

17. Brearton, C., et al., Performance Evaluation of a New Point of Care Viscoelastic Coagulation Monitoring System in Major Abdominal, Orthopaedic and Vascular Surgery. Platelets, 2020: p. 1–8.

18. Panigada, M., et al., Hypercoagulability of COVID-19 patients in intensive care unit: A report of thromboelastography findings and other parameters of hemostasis. J Thromb Haemost, 2020. 18(7): p. 1738–1742.

19. Balle, C.M., et al., Platelet Function During Extracorporeal Membrane Oxygenation in Adult Patients. Front Cardiovasc Med, 2019. 6: p. 114.

20. Meyer, A.D., et al., Platelet-derived microparticles generated by neonatal extracorporeal membrane oxygenation systems. ASAIO J, 2015. 61(1): p. 37–42.

